# Risk map of the novel coronavirus (2019-nCoV) in China: proportionate control is needed

**DOI:** 10.1101/2020.02.16.20023838

**Authors:** Xinhai Li, Xumao Zhao, Yingqiang Lou, Yuehua Sun

## Abstract

**Background:** China is running a national level antivirus campaign against the novel coronavirus (2019-nCoV). Strict control measures are being enforced in either the populated areas and remote regions. While the virus is closed to be under control, tremendous economic loss has been caused.

**Methods and findings:** We assessed the pandemic risk of 2019-nCoV for all cities/regions in China using the random forest algorithm, taking into account the effect of five factors: the accumulative and increased numbers of confirmed cases, total population, population density, and GDP. We defined four levels of the risk, corresponding to the four response levels to public health emergencies in China. The classification system has good consistency among cities in China, as the error rate of the confusion matrix is 1.58%.

**Conclusions:** The pandemic risk of 2019-nCoV is dramatically different among the 442 cities/regions. We recommend to adopt proportionate control policy according to the risk level to reduce unnecessary economic loss.

## 1. Introduction

The novel coronavirus (2019-nCoV) has infected 66,576 people and caused 1524 deaths in China by Feb. 15, 2020 [1]. It has spread to 25 countries [1] with the potential of further transmission [2-4]. On Jan. 30, 2020, the World Health Organization (WHO) declared the novel coronavirus outbreak to be “public health emergency of international concern” [5]. The countries with local infection of 2019-nCoV, such as Japan and Singapore, are implementing a moderate prevention policy against this flu-like infectious disease, similar to the situation of Wuhan in early January, 2020.

On the contrary, China is running a national level antivirus campaign. All of the 31 provinces and province-level municipalities in mainland China have initiated the first-level response to public health emergencies after the city closure of Wuhan on Jan. 23, 2020. Very strict control measures are enforced all over the country, such as tourism ban, school closure, meeting cancelation, etc. Compared with the prevention activities against SARS 17 years ago, traffic control and community locking are much more stricter, let alone the unprecedented province lockdown that constraining over 58 million people locally [6].

As a result, the control of 2019-nCoV in China is very effective [6-9]. The increases of the number of confirmed cases in most cities has dropped. No new cases have emerged in more than half cities in China on Feb. 14. While the half-lockout of industry, traffic, and school, etc., people are wondering how long the campaign will last and how soon the they can return to normal life.

We evaluated the risk of 2019-nCoV outbreak in cities and regions across China and suggested four levels of risk, highlighting that the cities with low risk should return to the normal status in order to avoid unnecessary economic loss.

## 2. Methods

### Data

We compiled the number of daily confirmed cases of 2019-nCoV (Fig.1) from the platform of real time epidemic situation for every city hosted by Baidu ™ at (https://voice.baidu.com/act/newpneumonia/newpneumonia/?from=osari_pc_1). We downloaded the data of population and gross domestic product (GDP) for every city in 2018 from the website of the National Bureau of Statistics of China at https://data.stats.gov.cn/easyquery.htm?cn=E0105 (Fig. 2A & 2C). The georeferenced data of the political boundaries of all prefecture level cities were provided by Institute of Geographic Sciences and Natural Resources Research, Chinese Academy of Sciences. The population density of the cities was calculated by dividing the population by the areas of the city (Fig. 2B).

**Fig 1.**
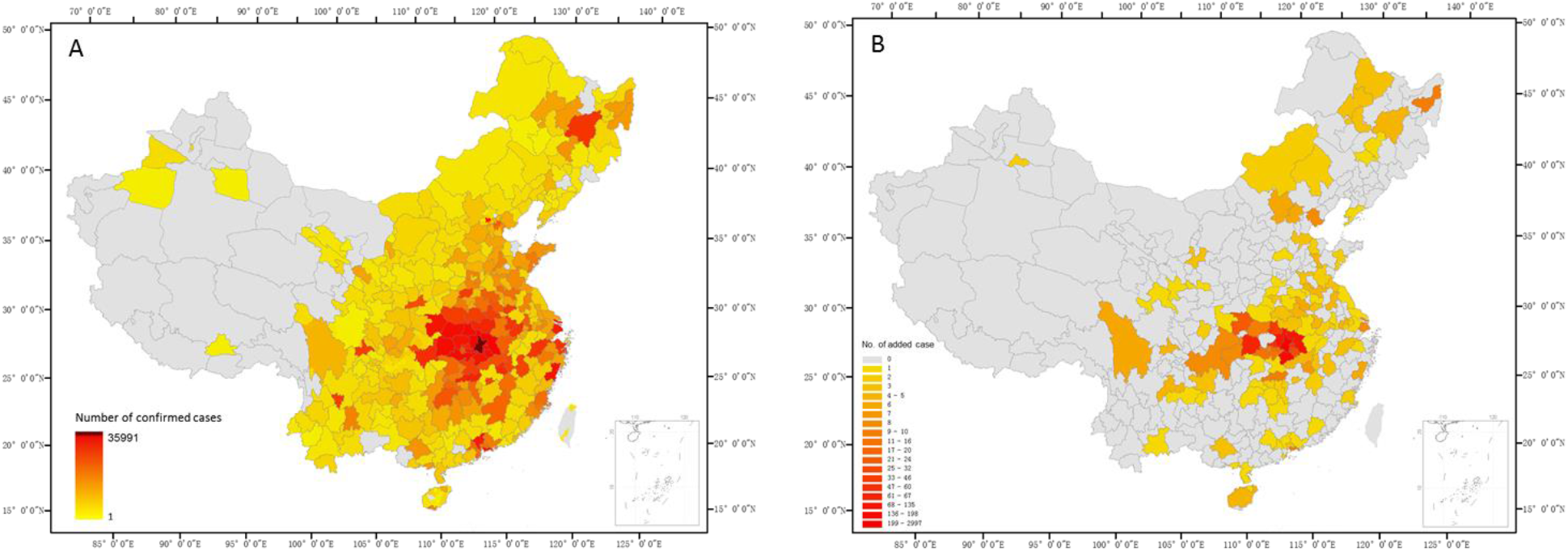
Number of cumulative confirmed cases (A) and increased cases (B) in cities across China on Feb. 14, 2020.

**Fig 2.**
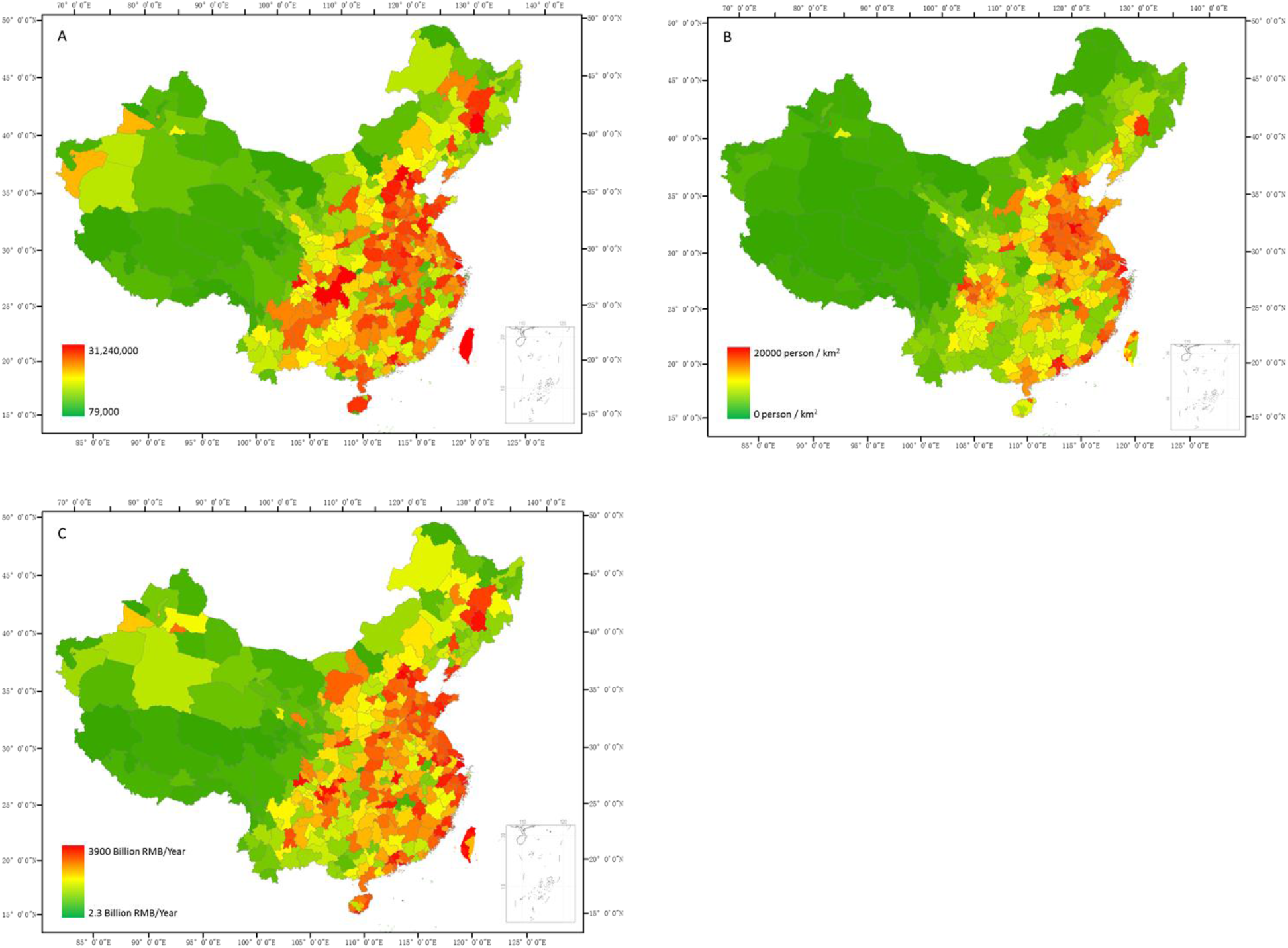
The population size (A), population density (B), and GDP (C) of the cities in China in 2018.

### Model

We used the random forest algorithm [10] and developed the following model:

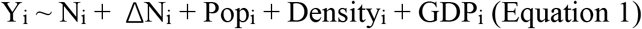

where *Y*_*i*_ is the response level of city *i* to public health emergencies; *N*_*i*_ is the number of cumulative confirmed cases on Feb. 14 in city *i*; *ΔN*_*i*_ is the increased number of cases on Feb. 14 in city *i*; *Pop*_*i*_, *Density*_*i*_, and *GDP*_*i*_ are the population size, population density, and GDP of city *i*, respectively.

The purpose of above model is to assign a response level, *Y*_*i*_ for each city. In the beginning, we manually assigned *Y*_*i*_ =1 for the top 10 cities with 2019-nCoV infection, and *Y*_*i*_ =4 for the 10 least infected cities. Then we gave random numbers for *Y*_*i*_ from {1, 2, 3, 4} to the rest of cities. We built the random forest model *RF* using Equation 1. *RF* contains the relationship between the dependent and independent variables. We used *RF* to predict the *Y*_*i*_ for every city, and used the newly calculated *Y*_*i*_ to update RF. We repeated the model learning process for 5-6 times until a low error rate was reached. In practice, we first defined *Y*_*i*_ as a continuous variable, quantifying the trend of *Y* using the five explanatory variables; and then defined *Y*_*i*_ as a categorical variable, classifying the response level of each city. At last, we manually assigned all cities (except for Shennongjia Forestry District, where has only 10 cases of 2019-nCoV) in Hubei Province with the highest category, the level I. The data and R code were posted at https://github.com/Xinhai-Li/2019-nCoV.

## 3. Results

Our model classified 442 cities with the four levels of response for public health emergencies (Fig. 3). There are 16 cities, all in Hubei Province, being assigned with the highest risk, the level I. The number of cities that were classified as risk levels II, III, and IV are 60, 67, and 299, respectively (Fig. 3).

**Fig 3.**
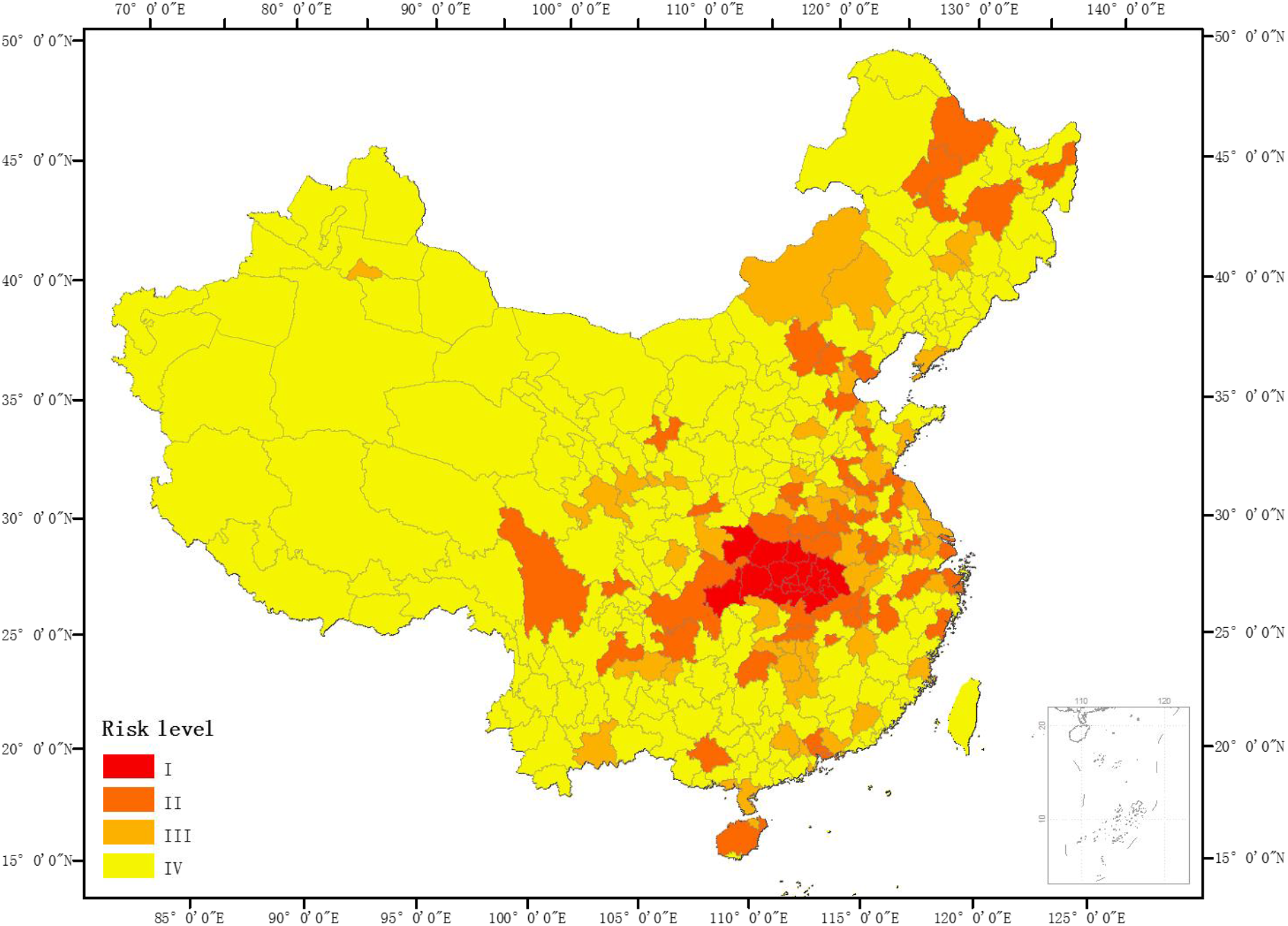
The risk levels of 2019-nCoV outbreak in cities/regions across China.

The classification for 442 cities has good consistency, as the overall error rate of the confusion matrix is only 1.58% (Table 1).

**Table 1.** The confusion matrix of the last random forest learning process. The response levels (1 to 4) was assigned to 442 cities.

| Previous prediction | Current prediction |  |  |  | Error |
| --- | --- | --- | --- | --- | --- |
|  | 1 | 2 | 3 | 4 |  |
| 1 | 11 | 4 | 0 | 1 | 31.3% |
| 2 | 0 | 59 | 0 | 1 | 1.7% |
| 3 | 0 | 1 | 66 | 0 | 1.5% |
| 4 | 0 | 0 | 0 | 299 | 0 |

The risk level of cities was classified using five variables, whereas the variables had different weights in the model, quantified by importance indices in random forest model [11] (Fig. 4).

**Fig 4.**
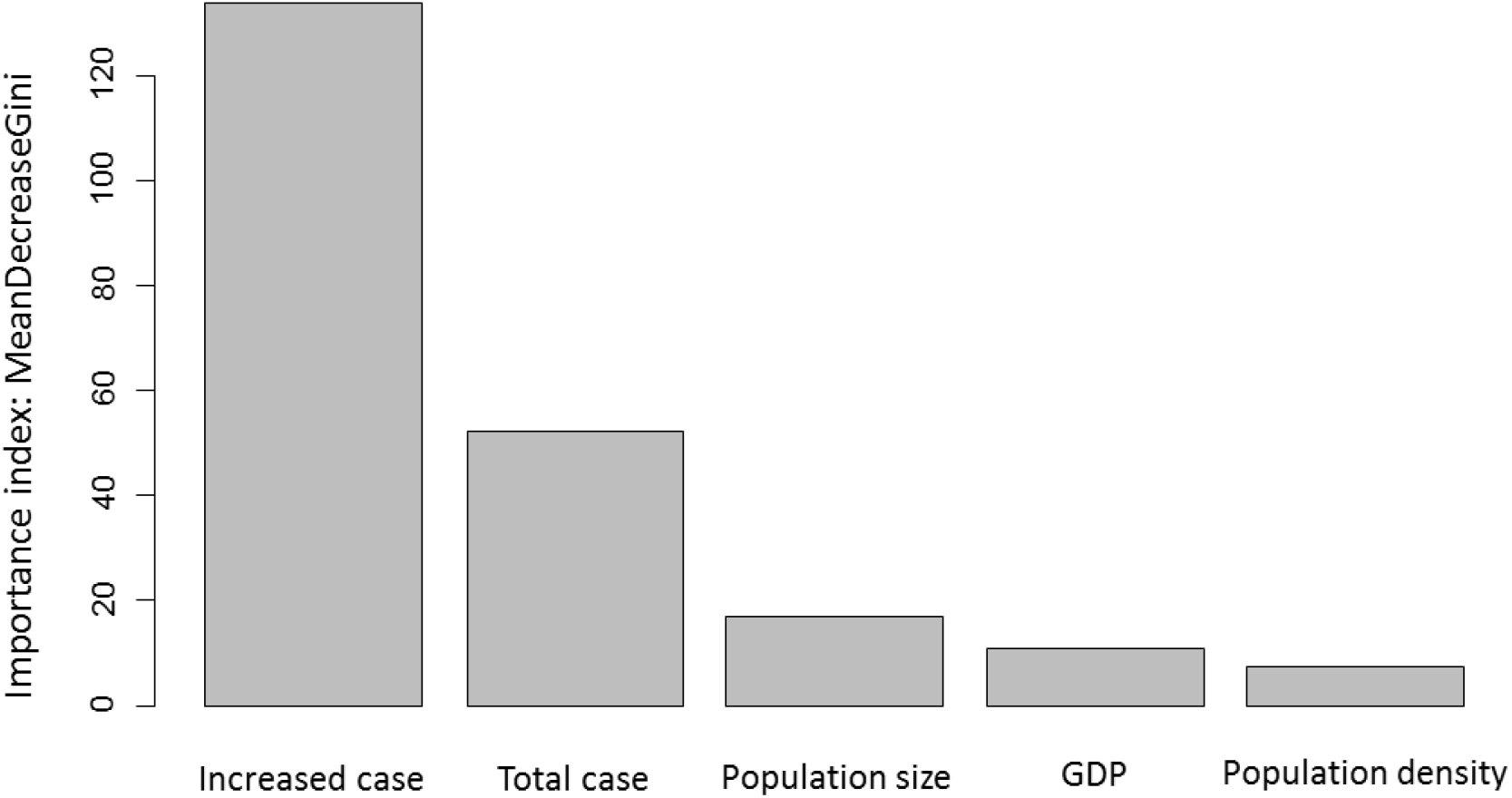
The importance of variables for classifying the risk levels of the 442 cities in China

## 4. Discussion

We assessed the risk level of 2019-nCoV outbreak for 442 cities in China based on five variables. The cumulative number of confirmed cases represents the size of the 2019-nCoV outbreak. The increased number of cases indicates the dynamics of virus transmission. Population size of a city is the key variable for epidemics, which is directly associated with the potential outbreak size. Population density is also an important factor, since dense population would accelerate virus transmission. Similarly, GDP is an index of economic activities, which links to human flow and virus transmission rate.

We used a machine learning algorithm, random forest, to classify the risk levels. One reason is that variables such as population size, population density and GDP are highly correlated. Random forest is very robust when multicollinearity is severe [12]. Another reason is that the numbers of cases are unbalanced among cities. In Wuhan, there are 39,249 confirmed cases by Feb. 16, whereas other large cities have a few hundred cases. Traditional linear models can not fit well with such a data distribution.

The purpose of our study is to provide a risk map of 2019-nCoV outbreak, so as to facilitate decision makers to adjust virus prevention policy. It is a great challenge to fight effectively and appropriately against a pandemic [13, 14], especially when we do not know much about the novel coronavirus [2, 15]. Rational and proportionate responses to such a virus outbreak are needed. At present, the strict control measures, for the areas with minor infection (which is under control), are overreacted.

The transmission of 2019-nCoV in cities outside Hubei Province is largely suspended [6]. From Feb.13 to Feb. 14, there are 231 cities have no new cases, and another 39 cities have only one new case (Fig. 1B). The risk in the Tibet Autonomous Region, Qinghai Province, Xinjiang Uygur Autonomous Region, Gansu Province, and Western Inner Mongolia Autonomous region is very low. We suggest rational, proportionate control measures (Table 2). For example, the cities with the risk level IV (Fig. 3) should return to normal status, under the condition of border control, in case of imported infections from high risk areas.

**Table 2.**
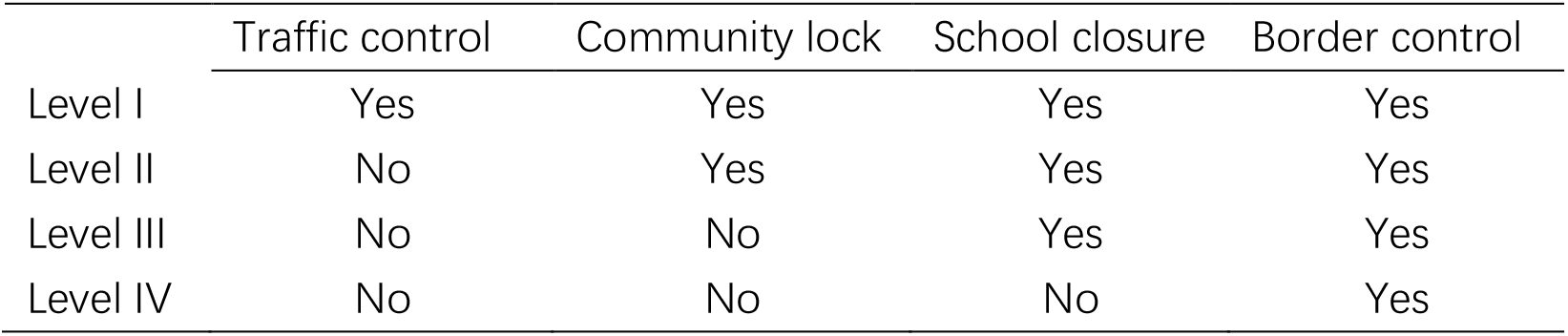
Suggested control measures following the four risk levels for every city

There is a high probability that a few transmission clusters of 2019-nCoV would happen after loosing the control in several provinces. Under such circumstance, the cities where virus returned should be managed again under higher level of responses to the public health emergencies.

Fighting against 2019-nCoV would be a long process, due to the three characteristics of this epidemics (long incubation period, asymptomatic infection, and high-false-negative-rate diagnosis) [6]. The transmission of 2019-nCoV is naturally (exponentially) growing in Japan and Singapore, increasing the risk of a global outbreak. Even after this transmission wave, 2019-nCoV might come back in every winter. As such, proportionate control is especially important for sustainable protect human from the infectious diseases.

## Data Availability

All data used in the study are free to public.

https://github.com/Xinhai-Li/2019-nCoV

## Acknowledgements

We are grateful to Professor Jon E. Swenson for help with English. We thank Yuqi Zou, Yongke Zhu, Tingting Yi, Chunlei Jing, Huan Liu, Boya Xie, Yiqing Li, Yumeng Tian for data processing.

